# MCA-UNet: A Multi-Scale Context and Attention U-Net for Colorectal Polyp Segmentation

**DOI:** 10.64898/2026.03.11.26348049

**Authors:** Yiqun Dong, Guhan Fang, Runsen Du, Haodong Hu, Zhengjie Fang, Chunhao Guo, Rongji Lu, Yuxin Jia, Ying Tian, Ziyi Wang

## Abstract

**Introduction:** To propose an improved U-Net-based segmentation model for colorectal polyp segmentation, aiming to address the challenges of variable lesion morphology, ambiguous boundaries, complex background interference, and insufficient cross-level feature fusion in endoscopic images [5,12].

**Methods:** An improved network termed MCA-UNet was developed based on U-Net [5]. The model incorporates a multi-scale context convolution block (MCCB) to enhance multi-scale feature extraction and an attention-guided feature fusion module (AGFF) to optimize skip-feature selection and fusion in the decoder. Experiments were conducted on publicly available colorectal polyp image datasets, including Kvasir-SEG and CVC-ClinicDB [13-15]. Four models, including U-Net, U-Net+MCCB, U-Net+AGFF, and MCA-UNet, were compared, and all models were trained for 100 epochs. Dice, intersection over union (IoU), and mean absolute error (MAE) were used as the main evaluation metrics [20].

**Results:** On the mixed validation set, the Dice scores of U-Net, U-Net+MCCB, U-Net+AGFF, and MCA-UNet were 0.742, 0.771, 0.754, and 0.783, respectively; the corresponding IoU values were 0.603, 0.635, 0.618, and 0.649; and the MAE values were 0.102, 0.090, 0.097, and 0.086. Compared with the baseline U-Net, MCA-UNet improved Dice and IoU by 5.53% and 7.63%, respectively, while reducing MAE by 15.69%. Comparisons on the Kvasir-SEG and CVC-ClinicDB validation subsets further demonstrated the more stable performance of the proposed model.

**Conclusion:** By jointly integrating multi-scale contextual modeling and attention-guided feature fusion, MCA-UNet effectively improves the accuracy and robustness of colorectal polyp segmentation and may provide useful support for intelligent endoscopic image analysis [12,17,18].

## 1. Introduction

Colorectal cancer is one of the most common malignant tumors of the digestive system, and its development is closely associated with the progression of colorectal polyps [1-3]. Therefore, early detection and accurate identification of colorectal polyps are of great clinical significance. Colonoscopy remains the primary tool for screening and diagnosing colorectal polyps and plays an essential role in colorectal cancer prevention [4]. Accurate lesion segmentation can further support lesion boundary localization, extent assessment, computer-aided diagnosis, and quantitative lesion analysis [12].

In recent years, convolutional neural network-based methods have achieved remarkable progress in medical image segmentation [5-10]. Among them, U-Net has been widely used in various medical image segmentation tasks due to its symmetric encoder-decoder architecture and skip connections [5]. In colorectal polyp segmentation, U-Net and its variants have also shown promising performance [12,16-19]. However, endoscopic polyp images are particularly challenging: polyps vary substantially in size, shape, texture, and color; some lesions have blurred boundaries and low contrast against surrounding mucosa; meanwhile, images are often affected by specular highlights, mucus, mucosal folds, and other complex background interference [12,14,15]. These factors limit the effectiveness of the standard U-Net in real-world applications.

On the one hand, conventional convolutions have limited receptive fields and cannot sufficiently model local details and broader contextual information at the same time [9]. On the other hand, in the decoder, the standard U-Net directly concatenates shallow and deep features through skip connections [5]. Although this operation helps recover spatial details, it may also introduce background noise and semantic mismatch across feature levels. Therefore, how to enhance both multi-scale contextual modeling and cross-level feature fusion while maintaining a relatively simple network structure remains a key issue in colorectal polyp segmentation [6,7,11].

To address these problems, we propose an improved colorectal polyp segmentation model termed MCA-UNet (Multi-scale Context and Attention U-Net). Built upon the U-Net framework [5], MCA-UNet introduces a multi-scale context convolution block (MCCB) to strengthen the joint modeling of local details and contextual information, and an attention-guided feature fusion module (AGFF) to optimize the selection and fusion of skip features. In this way, the proposed model aims to improve segmentation performance in complex endoscopic scenes.

The main contributions of this study are as follows. First, a multi-scale context convolution block, MCCB, is proposed to enhance multi-scale feature representation through parallel convolution branches with different receptive fields. Second, an attention-guided feature fusion module, AGFF, is introduced to refine shallow skip features before fusion and suppress irrelevant background responses, following the general idea of sequential channel-spatial attention refinement [11]. Third, MCA-UNet is constructed, and systematic experiments are conducted to validate the effectiveness of MCCB, AGFF, and their combination for colorectal polyp segmentation.

## 2. Materials and Methods

### 2.1 Overall Architecture of MCA-UNet

MCA-UNet is built upon the classical U-Net backbone [5] and adopts an encoder-decoder architecture. The network takes a three-channel RGB endoscopic image as input and outputs a single-channel binary segmentation map. Different from the standard U-Net, MCA-UNet introduces MCCB and AGFF into the feature extraction and feature fusion stages, respectively, to improve segmentation performance in complex-background and multi-scale lesion scenarios. The overall network architecture is illustrated in Figure 1.

**Figure 1.**
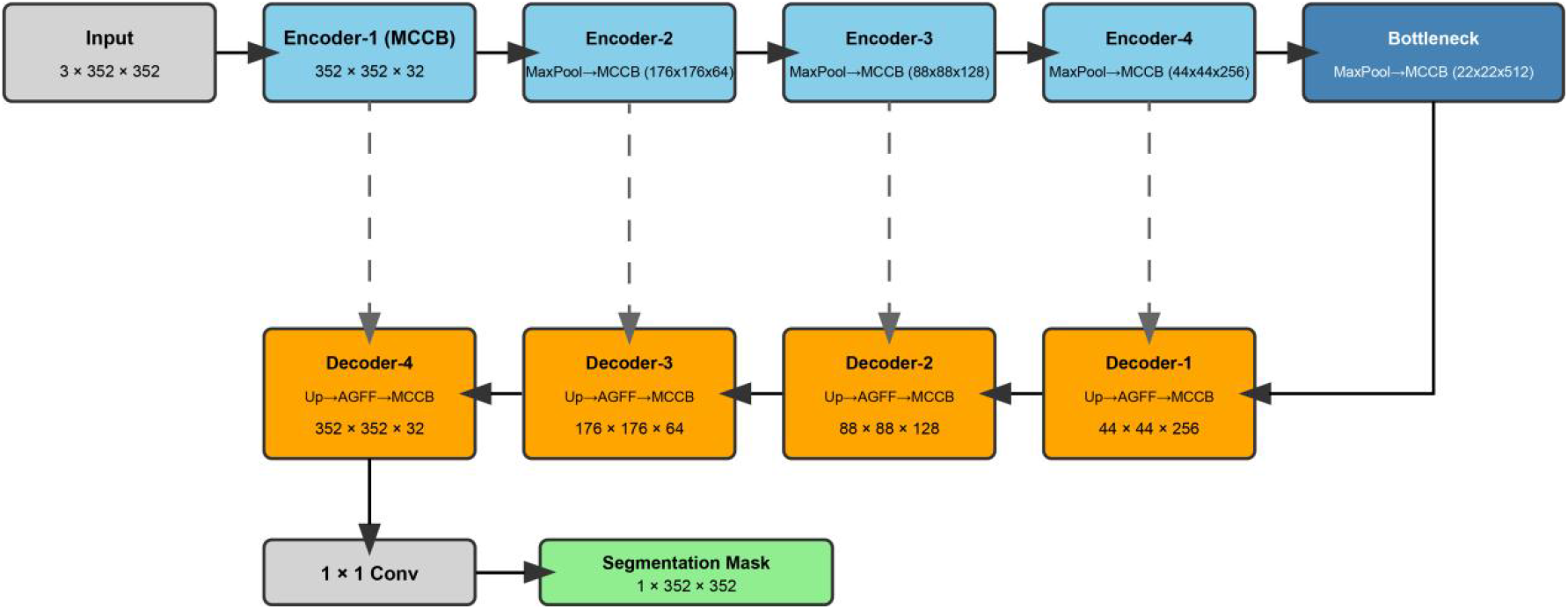
Overall architecture of MCA-UNet. The network is built upon the U-Net framework. MCCB is used in the encoder to enhance feature extraction, while the decoder adopts the sequence of upsampling, AGFF-based skip fusion, and MCCB-based refinement to reconstruct multi-scale features and generate the final polyp segmentation result.

In the encoder, conventional convolution blocks are replaced by MCCB to enhance feature extraction under different receptive fields. In the decoder, deep features are first upsampled, then combined with the corresponding encoder features through AGFF for skip-feature refinement, and finally processed by MCCB for multi-scale integration. Therefore, the core decoding process of MCA-UNet can be summarized as:

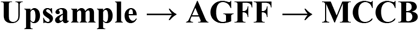

### 2.2 Baseline U-Net

To verify the effectiveness of the proposed modules, a standard U-Net was first constructed as the baseline model [5]. The model consists of an input convolution block, four encoder stages, four decoder stages, and an output layer. Each basic convolution block is implemented as a DoubleConv module, which contains two consecutive 3×3 convolutions, each followed by batch normalization and ReLU activation. In the encoder, max pooling is used to progressively reduce the spatial resolution and increase the channel dimension. In the decoder, bilinear interpolation is used for upsampling, followed by skip concatenation and convolutional refinement. Finally, a 1×1 convolution is used to generate single-channel segmentation logits.

### 2.3 Multi-Scale Context Convolution Block

The structure of the multi-scale context convolution block (MCCB) is shown in Figure 2. Different from the conventional DoubleConv block, MCCB adopts a parallel two-branch design. The first branch uses a standard 3×3 convolution to capture local texture and boundary details, while the second branch uses a 3×3 dilated convolution with dilation rate 2 to enlarge the receptive field and capture broader contextual information [9]. Both branches follow a convolution + batch normalization + ReLU configuration.

**Figure 2.**
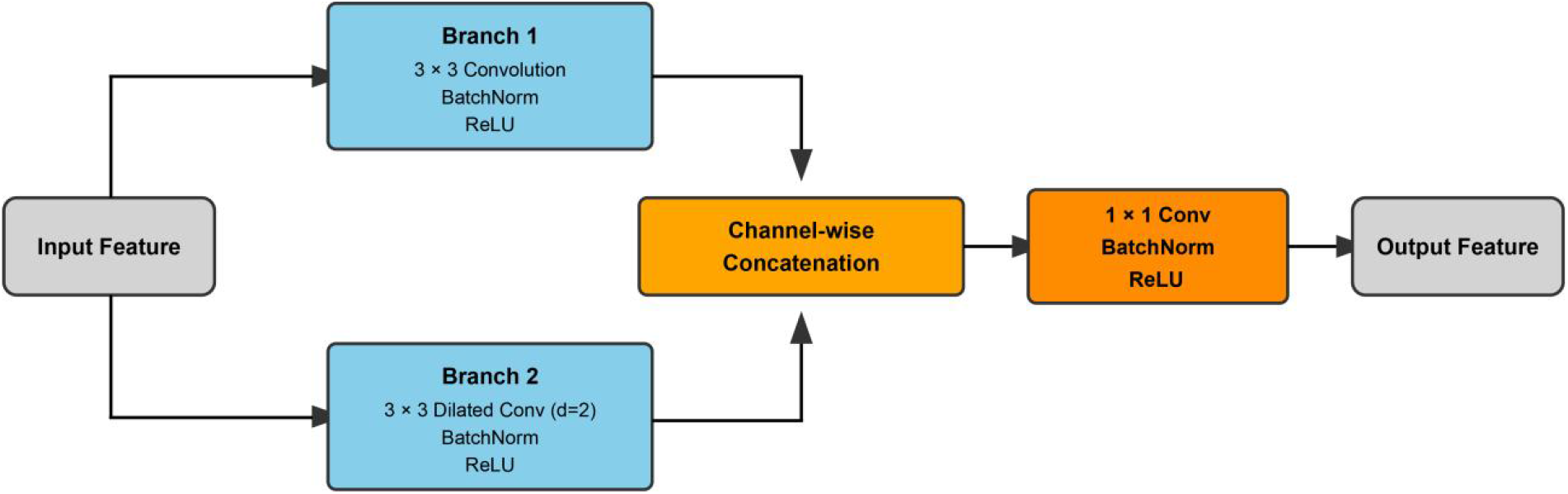
Structure of the multi-scale context convolution block (MCCB). The input feature is processed by a standard 3×3 convolution branch and a 3×3 dilated convolution branch with dilation rate 2 in parallel to extract local detail and contextual information, respectively. The outputs of the two branches are concatenated along the channel dimension and then fused by a 1×1 convolution.

The outputs of the two branches are concatenated along the channel dimension and then fused and compressed by a 1×1 convolution, followed by batch normalization and ReLU activation. This design enables the block to simultaneously model local details and contextual information.

### 2.4 Attention-Guided Feature Fusion Module

The structure of the attention-guided feature fusion module (AGFF) is shown in Figure 3. According to the actual implementation, the attention operations in AGFF are applied only to the skip feature rather than directly to the decoder feature. AGFF consists of two consecutive submodules: channel attention and spatial attention. This design is conceptually related to the sequential channel-spatial attention strategy adopted in CBAM [11].

**Figure 3.**
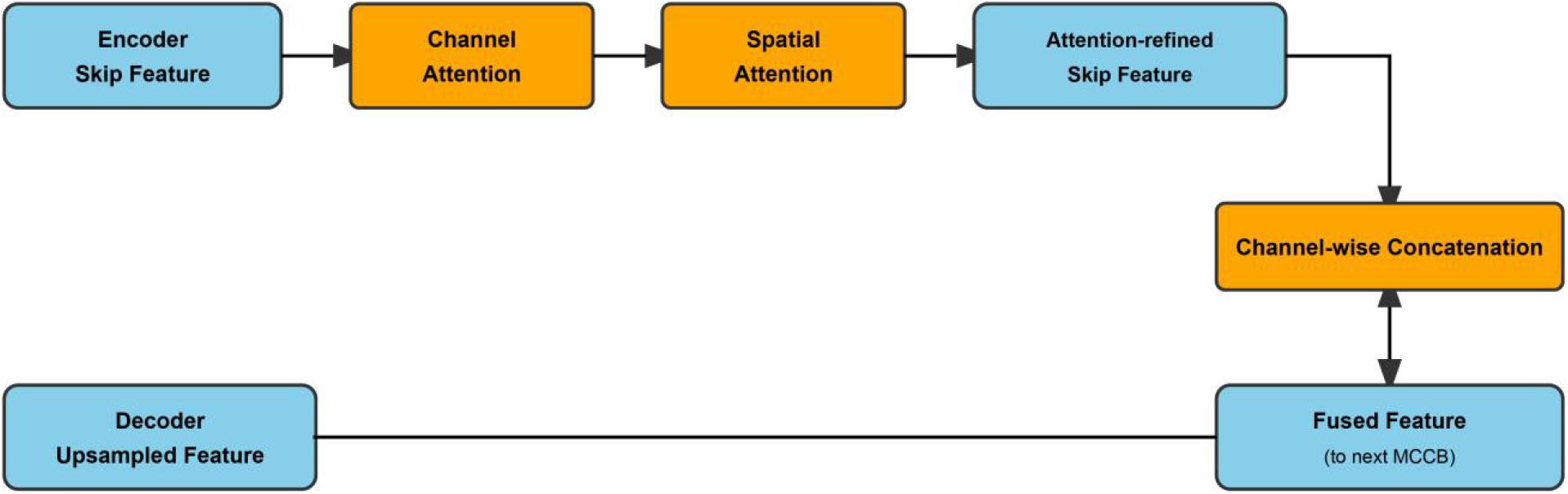
Structure of the attention-guided feature fusion module (AGFF). The skip feature from the encoder is sequentially refined by channel attention and spatial attention to highlight lesion-related responses and suppress background interference. The refined shallow feature is then concatenated with the upsampled decoder feature for subsequent feature integration.

In the channel attention stage, global average pooling is applied to the skip feature, and two 1×1 convolutions are used to generate channel-wise weights for feature recalibration [11]. In the spatial attention stage, average pooling and max pooling are performed along the channel dimension on the channel-refined feature, and the resulting maps are concatenated and passed through a 7×7 convolution followed by sigmoid activation to generate a spatial attention map [11]. This map is multiplied element-wise with the input feature to highlight lesion-related regions and suppress irrelevant background responses.

Finally, the refined skip feature is concatenated with the upsampled decoder feature along the channel dimension to form the output of AGFF.

### 2.5 Decoder Design

In U-Net+AGFF, the encoder keeps the standard DoubleConv structure, while the decoder adopts the following design:

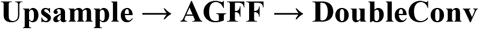

In MCA-UNet, both encoder and decoder convolution blocks are replaced by MCCB, and the decoder follows:

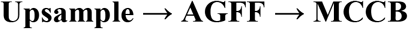

This design allows AGFF to address the problem of which shallow features should be fused, while MCCB focuses on enhancing the fused features through multi-scale contextual modeling [9,11].

### 2.6 Compared Models and Experimental Design

To systematically evaluate the independent and joint contributions of the proposed modules, four models were constructed for comparison:

1. *U-Net: baseline model [5];*
2. *U-Net+MCCB: replacing the standard convolution blocks with MCCB only;*
3. *U-Net+AGFF: introducing AGFF into the decoder only;*
4. *MCA-UNet: incorporating both MCCB and AGFF*.

In addition, to further analyze the contribution of different attention components in AGFF, an ablation experiment was conducted by comparing channel attention only, spatial attention only, and the combination of both [11].

### 2.7 Dataset Composition, Preprocessing, and Augmentation

Publicly available colorectal polyp image datasets, namely Kvasir-SEG [13] and CVC-ClinicDB [14,15], were used in this study. For unified training and evaluation, the two datasets were first split into training and validation subsets separately, and then the corresponding subsets were merged to form a mixed training set and a mixed validation set. Model training was performed on the mixed training set, while the main results were calculated on the mixed validation set. Additional evaluations were conducted separately on the Kvasir-SEG validation subset and the CVC-ClinicDB validation subset to analyze model performance across different data sources.

The overall metrics on the mixed validation set were calculated using an image-wise averaging strategy. Specifically, Dice, IoU, and MAE were first computed for each validation image and then averaged over all validation images. The same strategy was applied to the Kvasir-SEG and CVC-ClinicDB validation subsets. Therefore, the overall results were directly computed from the mixed validation set rather than simply inferred by weighted averaging of the two subset results.

Input images and corresponding masks were stored in “images” and “masks” folders, respectively, and matched by filename stem. Multiple image formats were supported, including .png, .jpg, .jpeg, .bmp, .tif, and .tiff. Masks were read as grayscale images and converted into binary labels by thresholding.

All images were resized to 352×352 before being fed into the network. Online data augmentation was applied during training, including random scaling, random rotation, horizontal flipping, and vertical flipping. The scaling range was set to −0.25 to 0.25, and the rotation angle was limited to ±30°. No random augmentation was applied during validation; only resizing and normalization were performed. The normalization setting was equivalent to scaling the original pixel intensities to the range [0, 1].

### 2.8 Training Details and Loss Function

The models were implemented in PyTorch. Training data were fed into the network through a DataLoader. AdamW was used as the optimizer, with an initial learning rate of 1×10^-4 and a weight decay of 1×10^-4 [21]. All models were trained for 100 epochs to ensure fair comparison. The input image size was 352×352, and the base channel number was set to 32. All experiments were conducted on an NVIDIA GeForce RTX 3090 GPU.

A hybrid loss function combining BCEWithLogitsLoss and DiceLoss was adopted, with equal weights:

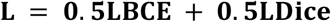

where DiceLoss is defined as:

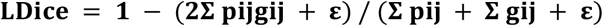

where pij denotes the predicted probability after sigmoid activation, gij denotes the ground-truth label, and ε = 10^−6. Dice-based loss functions are widely used in medical image segmentation, especially for handling foreground-background imbalance [10,20].

### 2.9 Evaluation Metrics

Dice, IoU, and MAE were used as the main evaluation metrics [10,20]. The network output was first converted into a probability map by sigmoid activation and then binarized using a threshold of 0.5.

Dice is defined as:

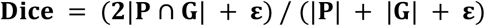

IoU is defined as:

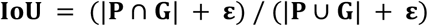

MAE is defined as:

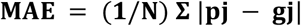

where P and G denote the predicted foreground and ground-truth foreground, respectively, and N denotes the total number of pixels.

## 3. Results

### 3.1 Overall Comparison on the Mixed Validation Set

The overall segmentation results of different models on the mixed validation set are shown in Table 1. The baseline U-Net achieved a Dice of 0.742, an IoU of 0.603, and an MAE of 0.102. After introducing MCCB, the corresponding metrics improved to 0.771, 0.635, and 0.090, respectively. After introducing AGFF, the corresponding metrics became 0.754, 0.618, and 0.097. Meanwhile, MCA-UNet, which incorporates both MCCB and AGFF, achieved the best overall performance, with a Dice of 0.783, an IoU of 0.649, and an MAE of 0.086.

**Table 1.**
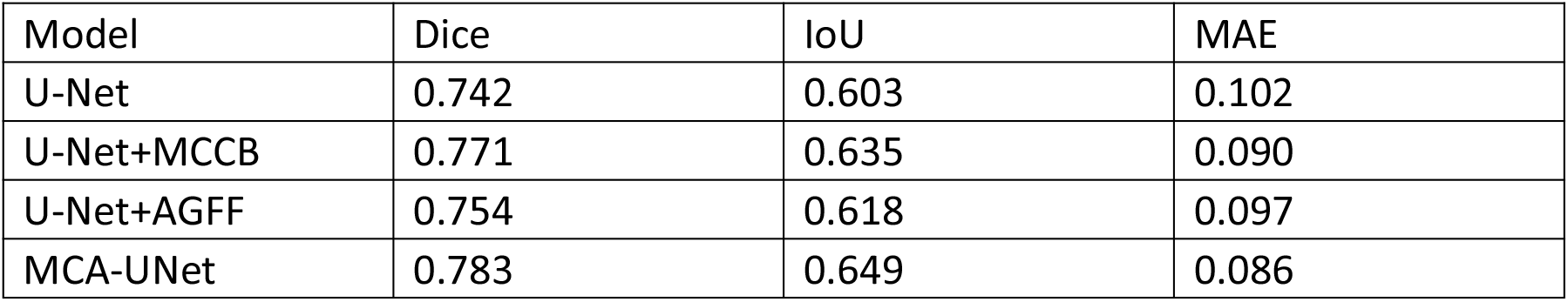
Overall comparison of different models on the mixed validation set.

| Model | Dice | IoU | MAE |
| --- | --- | --- | --- |
| U-Net | 0.742 | 0.603 | 0.102 |
| U-Net+MCCB | 0.771 | 0.635 | 0.090 |
| U-Net+AGFF | 0.754 | 0.618 | 0.097 |
| MCA-UNet | 0.783 | 0.649 | 0.086 |

Overall, both MCCB and AGFF improved the segmentation performance of the baseline model, with MCCB providing a more pronounced gain. When combined, the two modules led to further improvement, indicating that multi-scale contextual modeling and attention-guided feature fusion are complementary for colorectal polyp segmentation.

### 3.2 Ablation Analysis

To evaluate the independent and joint contributions of MCCB and AGFF, ablation experiments were performed, and the results are shown in Table 2. Compared with the baseline U-Net, introducing MCCB alone improved Dice and IoU by 3.91% and 5.31%, respectively, while reducing MAE by 11.76%. Introducing AGFF alone improved Dice and IoU by 1.62% and 2.49%, respectively, while reducing MAE by 4.90%. When both MCCB and AGFF were incorporated, MCA-UNet achieved the best performance, with Dice and IoU improved by 5.53% and 7.63%, respectively, and MAE reduced by 15.69%. These results indicate that both MCCB and AGFF contribute to performance improvement, and their combination yields a more evident synergistic gain.

**Table 2.**
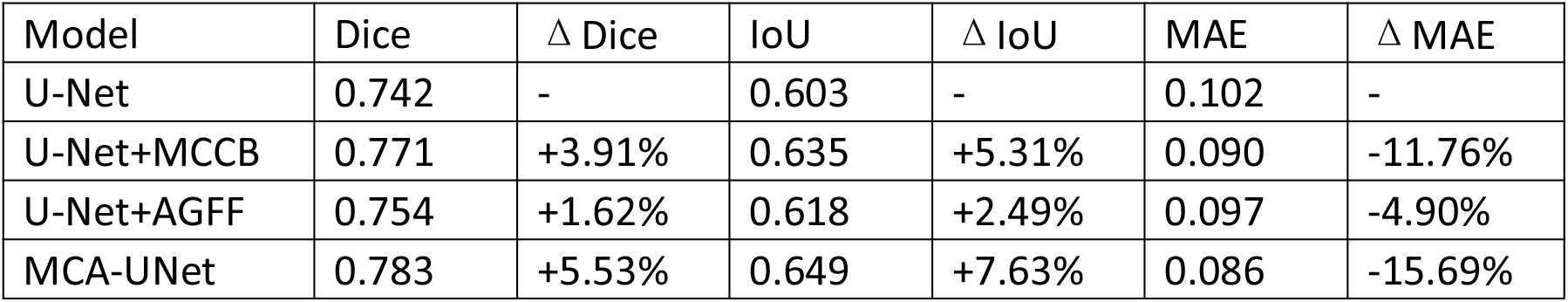
Ablation results of MCCB and AGFF and their relative improvements over the baseline.

| Model | Dice | $\Delta$ Dice | IoU | $\Delta$ IoU | MAE | $\Delta$ MAE |
| --- | --- | --- | --- | --- | --- | --- |
| U-Net | 0.742 | - | 0.603 | - | 0.102 | - |
| U-Net+MCCB | 0.771 | +3.91% | 0.635 | +5.31% | 0.090 | -11.76% |
| U-Net+AGFF | 0.754 | +1.62% | 0.618 | +2.49% | 0.097 | -4.90% |
| MCA-UNet | 0.783 | +5.53% | 0.649 | +7.63% | 0.086 | -15.69% |

### 3.3 Analysis of AGFF Components

To further investigate the contribution of different attention components in AGFF, we compared channel attention only, spatial attention only, and the combination of both. The results are presented in Table 3. Both channel attention and spatial attention individually improved the baseline U-Net, with channel attention providing a slightly larger gain. Meanwhile, the full U-Net+AGFF model, which combines both channel and spatial attention, achieved the best performance, indicating that the dual-attention design in AGFF more effectively refines skip features and improves fusion quality [11].

**Table 3.** Ablation results of different AGFF components.

| Variant | Channel Attention | Spatial Attention | Dice | IoU | MAE |
| --- | --- | --- | --- | --- | --- |
| U-Net | - | - | 0.742 | 0.603 | 0.102 |
| U-Net+CA | ✓ | - | 0.752 | 0.615 | 0.098 |
| U-Net+SA | - | ✓ | 0.748 | 0.610 | 0.100 |
| U-Net+AGFF | ✓ | ✓ | 0.754 | 0.618 | 0.097 |

### 3.4 Comparison on Different Validation Subsets

To further evaluate model performance across different data sources, we compared all models on the Kvasir-SEG validation subset and the CVC-ClinicDB validation subset, respectively, and the results are shown in Table 4 and Table 5.

**Table 4.** Comparison of different models on the Kvasir-SEG validation subset.

| Model | Dice | IoU | MAE |
| --- | --- | --- | --- |
| U-Net | 0.769 | 0.636 | 0.094 |
| U-Net+MCCB | 0.801 | 0.671 | 0.081 |
| U-Net+AGFF | 0.780 | 0.649 | 0.089 |
| MCA-UNet | 0.808 | 0.680 | 0.078 |

**Table 5.** Comparison of different models on the CVC-ClinicDB validation subset.

| Model | Dice | IoU | MAE |
| --- | --- | --- | --- |
| U-Net | 0.713 | 0.569 | 0.109 |
| U-Net+MCCB | 0.741 | 0.601 | 0.097 |
| U-Net+AGFF | 0.733 | 0.589 | 0.101 |
| MCA-UNet | 0.758 | 0.621 | 0.091 |

On the Kvasir-SEG validation subset, the Dice scores of U-Net, U-Net+MCCB, U-Net+AGFF, and MCA-UNet were 0.769, 0.801, 0.780, and 0.808, respectively; the corresponding IoU values were 0.636, 0.671, 0.649, and 0.680; and the MAE values were 0.094, 0.081, 0.089, and 0.078. MCA-UNet achieved the best performance on this subset.

On the CVC-ClinicDB validation subset, the overall performance of all models was lower than that on the Kvasir-SEG validation subset. The Dice scores of U-Net, U-Net+MCCB, U-Net+AGFF, and MCA-UNet were 0.713, 0.741, 0.733, and 0.758, respectively; the corresponding IoU values were 0.569, 0.601, 0.589, and 0.621; and the MAE values were 0.109, 0.097, 0.101, and 0.091. MCA-UNet again achieved the best performance.

Overall, MCA-UNet maintained relatively stable superiority on both validation subsets, suggesting that the proposed model has a certain degree of robustness across different visual distributions [13-15].

### 3.5 Model Complexity and Inference Efficiency

The model complexity and inference efficiency are summarized in Table 6. Compared with the baseline U-Net, introducing MCCB or AGFF increased the number of parameters, computational cost, and inference time per image. MCCB had a slightly greater impact on model complexity than AGFF, while MCA-UNet, which incorporates both modules, showed the highest parameter count and FLOPs. Nevertheless, MCA-UNet achieved the best Dice and IoU scores and the lowest MAE, suggesting that it obtained better segmentation performance with an acceptable increase in complexity.

**Table 6.** Comparison of model complexity and inference efficiency.

| Model | Params(M) | FLOPs(G) | Time(ms/image) | Dice | IoU | MAE |
| --- | --- | --- | --- | --- | --- | --- |
| U-Net | 7.76 | 16.42 | 14.8 | 0.742 | 0.603 | 0.102 |
| U-Net+MCCB | 8.34 | 18.15 | 16.3 | 0.771 | 0.635 | 0.090 |
| U-Net+AGFF | 7.92 | 16.88 | 15.6 | 0.754 | 0.618 | 0.097 |
| MCA-UNet | 8.57 | 18.74 | 17.2 | 0.783 | 0.649 | 0.086 |

Table 6. Comparison of model complexity and inference efficiency

### 3.6 Qualitative Visualization Analysis

Representative qualitative segmentation results are shown in Figure 4. The baseline U-Net tended to produce local under-segmentation, discontinuous boundaries, and false positives under complex background conditions or when lesions were small or had blurred boundaries, which is consistent with the known challenges of colorectal polyp segmentation [12,14,15]. After introducing MCCB, the model improved its ability to capture the overall contour of polyps; after introducing AGFF, false segmentation in complex background regions was reduced. In contrast, MCA-UNet produced segmentation results closer to the ground truth in most cases, with better lesion completeness, boundary continuity, and detail recovery.

**Figure 4.**
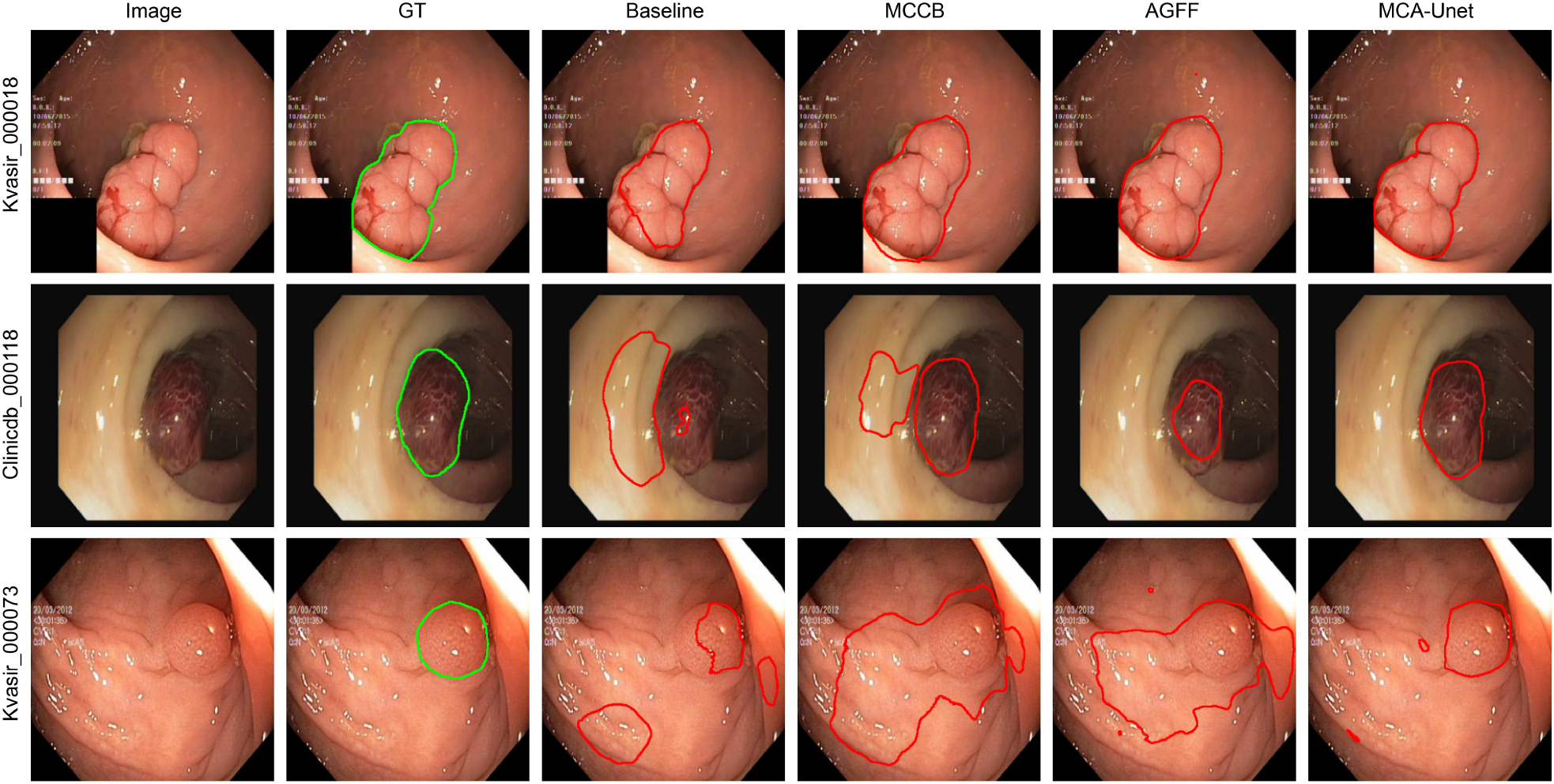
Qualitative comparison of segmentation results produced by different models on representative colorectal polyp images. From left to right are the input image, ground truth, U-Net, U-Net+MCCB, U-Net+AGFF, and MCA-UNet. Compared with the competing models, MCA-UNet achieves better lesion completeness, boundary continuity, and suppression of false segmentation under complex background conditions.

## 4. Discussion

This study proposed MCA-UNet, an improved model for colorectal polyp segmentation. Based on U-Net [5], the proposed model introduces targeted improvements in two key aspects: feature extraction and feature fusion. MCCB is used to enhance multi-scale contextual modeling, while AGFF is designed to improve the quality of skip-feature fusion. Compared with simply increasing network depth or width, the proposed strategy focuses more directly on the core challenges of the task [9,11].

The experimental results showed that both MCCB and AGFF improved the segmentation performance of the baseline U-Net, while their combination achieved better overall performance. This indicates that the improvement in colorectal polyp segmentation is not due to a single module alone, but rather to the combined effects of enhanced multi-scale representation and optimized cross-level fusion. MCCB alone produced a larger gain than AGFF, suggesting that multi-scale feature extraction may be a more direct factor influencing performance in this task. Meanwhile, although AGFF alone yielded a relatively smaller gain, it still provided further improvement when combined with MCCB, indicating that its refinement effect in feature fusion is independently valuable. The AGFF component analysis further showed that both channel attention and spatial attention contributed to performance improvement, and their combination was the most effective, supporting the rationality of the AGFF design [11].

The complexity analysis showed that MCA-UNet moderately increased the number of parameters, FLOPs, and inference time compared with the baseline U-Net. However, this increase remained acceptable in view of the observed performance gain. Therefore, the proposed model achieved a relatively good balance between segmentation accuracy and computational efficiency. Compared with some recent polyp segmentation architectures, such as ResUNet++, PraNet, and HarDNet-MSEG [16-18], the present study places more emphasis on modular improvement based on a clear U-Net framework rather than introducing a substantially more complex overall design.

This study still has several limitations. First, the experiments were conducted mainly on publicly available datasets [13-15], and the data scale and imaging scenarios remain limited. Thus, the cross-dataset generalization ability of the model still needs to be further validated on more external datasets. Second, this study mainly adopted Dice, IoU, and MAE as evaluation metrics [10,20], while additional metrics widely used in polyp segmentation could be incorporated in future work for more comprehensive evaluation. Third, this work focused mainly on model design and segmentation performance, while further interpretability analyses, such as attention distribution and feature response visualization, remain to be explored.

## 5. Conclusion

In this study, an improved U-Net-based model, MCA-UNet, was proposed for colorectal polyp segmentation. By introducing the multi-scale context convolution block MCCB and the attention-guided feature fusion module AGFF, the proposed model enhanced both feature representation and decoder fusion, thereby achieving collaborative improvement in feature extraction and feature recovery.

Experimental results showed that MCA-UNet achieved better performance on the mixed validation set as well as on different source-specific validation subsets. The proposed model demonstrated promising capability in lesion identification, boundary recovery, and suppression of complex background interference. Overall, MCA-UNet provides a structurally clear, logically complete, and practically valuable solution for colorectal polyp segmentation [12,17,18].

## Acknowledgments

The authors thank the developers of the Kvasir-SEG and CVC-ClinicDB datasets for making these public datasets available [13-15].

## CRediT Author Statement

Yiqun Dong: Conceptualization, Supervision, Writing – review and editing, Correspondence.

Guhan Fang: Methodology, Software, Formal analysis, Visualization, Writing – original draft.

Runsen Du: Software, Validation, Formal analysis, Writing – review and editing. Haodong Hu, Zhengjie Fang, Chunhao Guo, Rongji Lu, Yuxin Jia, Yin Tian, Ziyi Wang: Investigation, Data curation, Validation, Writing – review and editing.

Yiqun Dong, Guhan Fang, and Runsen Du contributed equally to this work and share first authorship.

## Data Availability Statement

The datasets used in this study are publicly available, including Kvasir-SEG [13] and CVC-ClinicDB [14,15]. The source code is publicly available at: https://github.com/DYQ04/colorectal-polyps

## Ethics Statement

This study was based exclusively on publicly available anonymized datasets [13-15]. Ethical approval and informed consent were not required.

## Declaration of Generative AI and AI-Assisted Technologies in the Manuscript Preparation Process

During the preparation of this manuscript, generative AI tools were used to assist with language polishing and text refinement. All content was reviewed by the authors, who take full responsibility for the final manuscript.

## References

[1] Bray F, Laversanne M, Sung H, Ferlay J, Siegel RL, Soerjomataram I, et al. Global cancer statistics 2022: GLOBOCAN estimates of incidence and mortality worldwide for 36 cancers in 185 countries. CA Cancer J Clin. 2024;74(3):229–263.

[2] Siegel RL, Giaquinto AN, Jemal A. Cancer statistics, 2024. CA Cancer J Clin. 2024;74(1):12–49.

[3] Øines M, Helsingen LM, Bretthauer M, Emilsson L. Epidemiology and risk factors of colorectal polyps. Best Pract Res Clin Gastroenterol. 2017;31(4):419–424.

[4] Helsingen LM, Kalager M. Colorectal cancer screening—approach, evidence, and future directions. NEJM Evid. 2022;1(1):EVIDra2100035.

[5] Ronneberger O, Fischer P, Brox T. U-Net: Convolutional Networks for Biomedical Image Segmentation. In: Medical Image Computing and Computer-Assisted Intervention – MICCAI 2015. Cham: Springer; 2015. p. 234–241.

[6] Zhou Z, Siddiquee MMR, Tajbakhsh N, Liang J. UNet++: A nested U-Net architecture for medical image segmentation. In: Deep Learning in Medical Image Analysis and Multimodal Learning for Clinical Decision Support. Cham: Springer; 2018. p. 3–11.

[7] Oktay O, Schlemper J, Le Folgoc L, Lee M, Heinrich M, Misawa K, et al. Attention U-Net: Learning where to look for the pancreas. arXiv. 2018:1804.03999.

[8] Badrinarayanan V, Kendall A, Cipolla R. SegNet: A deep convolutional encoder-decoder architecture for image segmentation. IEEE Trans Pattern Anal Mach Intell. 2017;39(12):2481–2495.

[9] Chen LC, Zhu Y, Papandreou G, Schroff F, Adam H. Encoder-Decoder with Atrous Separable Convolution for Semantic Image Segmentation. In: Proceedings of the European Conference on Computer Vision (ECCV). 2018. p. 801–818.

[10] Milletari F, Navab N, Ahmadi SA. V-Net: Fully convolutional neural networks for volumetric medical image segmentation. In: 2016 Fourth International Conference on 3D Vision (3DV). 2016. p. 565–571.

[11] Woo S, Park J, Lee JY, Kweon IS. CBAM: Convolutional Block Attention Module. In: Computer Vision – ECCV 2018. Cham: Springer; 2018. p. 3–19.

[12] Sánchez-Peralta LF, Bote-Curiel L, Picón A, Sánchez-Margallo FM, Pagador JB. Deep learning to find colorectal polyps in colonoscopy: A systematic literature review. Artif Intell Med. 2020;108:101923.

[13] Jha D, Smedsrud PH, Riegler MA, Halvorsen P, de Lange T, Johansen D, et al. Kvasir-SEG: A segmented polyp dataset. In: MultiMedia Modeling. Cham: Springer; 2020. p. 451–462.

[14] Bernal J, Sánchez FJ, Fernández-Esparrach G, Gil D, Rodríguez C, Vilariño F. WM-DOVA maps for accurate polyp highlighting in colonoscopy: Validation vs. saliency maps from physicians. Comput Med Imaging Graph. 2015;43:99–111.

[15] Bernal J, Sánchez J, Vilariño F. Comparative validation of polyp detection methods in video colonoscopy: Results from the MICCAI 2015 Endoscopic Vision Challenge. IEEE Trans Med Imaging. 2017;36(6):1231–1249.

[16] Jha D, Smedsrud PH, Riegler MA, Halvorsen P, de Lange T, Johansen D, et al. ResUNet++: An advanced architecture for medical image segmentation. arXiv. 2019:1911.07067.

[17] Fan DP, Ji GP, Zhou T, Chen G, Fu H, Shen J, et al. PraNet: Parallel Reverse Attention Network for Polyp Segmentation. In: Medical Image Computing and Computer Assisted Intervention – MICCAI 2020. Cham: Springer; 2020. p. 263–273.

[18] Huang CH, Wu HY, Lin YL. HarDNet-MSEG: A simple encoder-decoder polyp segmentation neural network that achieves over 0.9 mean Dice and 86 FPS. arXiv. 2021:2101.07172.

[19] Alom MZ, Hasan M, Yakopcic C, Taha TM, Asari VK. Recurrent residual U-Net for medical image segmentation. J Med Imaging. 2019;6(1):014006.

[20] Sudre CH, Li W, Vercauteren T, Ourselin S, Cardoso MJ. Generalised Dice overlap as a deep learning loss function for highly unbalanced segmentations. In: Deep Learning in Medical Image Analysis and Multimodal Learning for Clinical Decision Support. Cham: Springer; 2017. p. 240–248.

[21] Loshchilov I, Hutter F. Decoupled weight decay regularization. In: International Conference on Learning Representations (ICLR). 2019.

